# Risk Prediction of Coronary Artery Spasm in Patients Without Obstructive Coronary Artery Disease Using a Comprehensive Clinical, Laboratory and Echocardiographic Diagnostic Score

**DOI:** 10.1101/2025.10.01.25337133

**Authors:** Yu-Ching Lee, Ian Y Chen, Ming-Jui Hung, Chi-Tai Yeh, Nicholas G. Kounis, Patrick Hu, Ming-Yow Hung

## Abstract

**BACKGROUND:** The lack of an accurate coronary artery spasm (CAS) risk prediction model highlights the failure to consider the dynamic coronary health and a gap in understanding CAS.

**METHODS:** A total of 913 Taiwanese patients (460 women and 453 men) with suspected ischemic heart disease but without angiographic obstructive coronary artery disease were subjected to intracoronary methylergonovine testing during the period 2008-2025.

**RESULTS:** The study included 645 CAS cases (70.6%) and 268 non-CAS controls (29.4%). The proportion of men was higher in the CAS than non-CAS group (54.6% vs. 37.7%, p<0.001). The multivariable logistic regression model identified 10 variables significantly associated with CAS (p<0.05): male, smoking, low systolic and diastolic blood pressure, reduced B-type natriuretic peptide levels, elevated low-density lipoprotein levels, increased relative wall thickness at end-systole, high left ventricular mass index, low e’(l) values and high Tei index. Notably, concentric remodeling might promote CAS development. Discrimination performance was moderate, with an AUC value of 73.8% that dropped to 72.4% after bootstrapped internal validation, suggesting the potential generalizability of the derived model. The scoring system ranged from 36 to 98, representing predicted probabilities between 12% and 98%, respectively.

**CONCLUSIONS:** While a total score of ≥58 with the probability of CAS exceeding 50% indicates a significant chance of undiagnosed CAS, for those with a total score ≥69 with a very high probability of CAS ≥75%, coronary catheterization with CAS provocation tests is strongly recommended for a definite diagnosis. The simple 10-variable scoring model allows ranking at-risk population and diagnostic resource allocation.

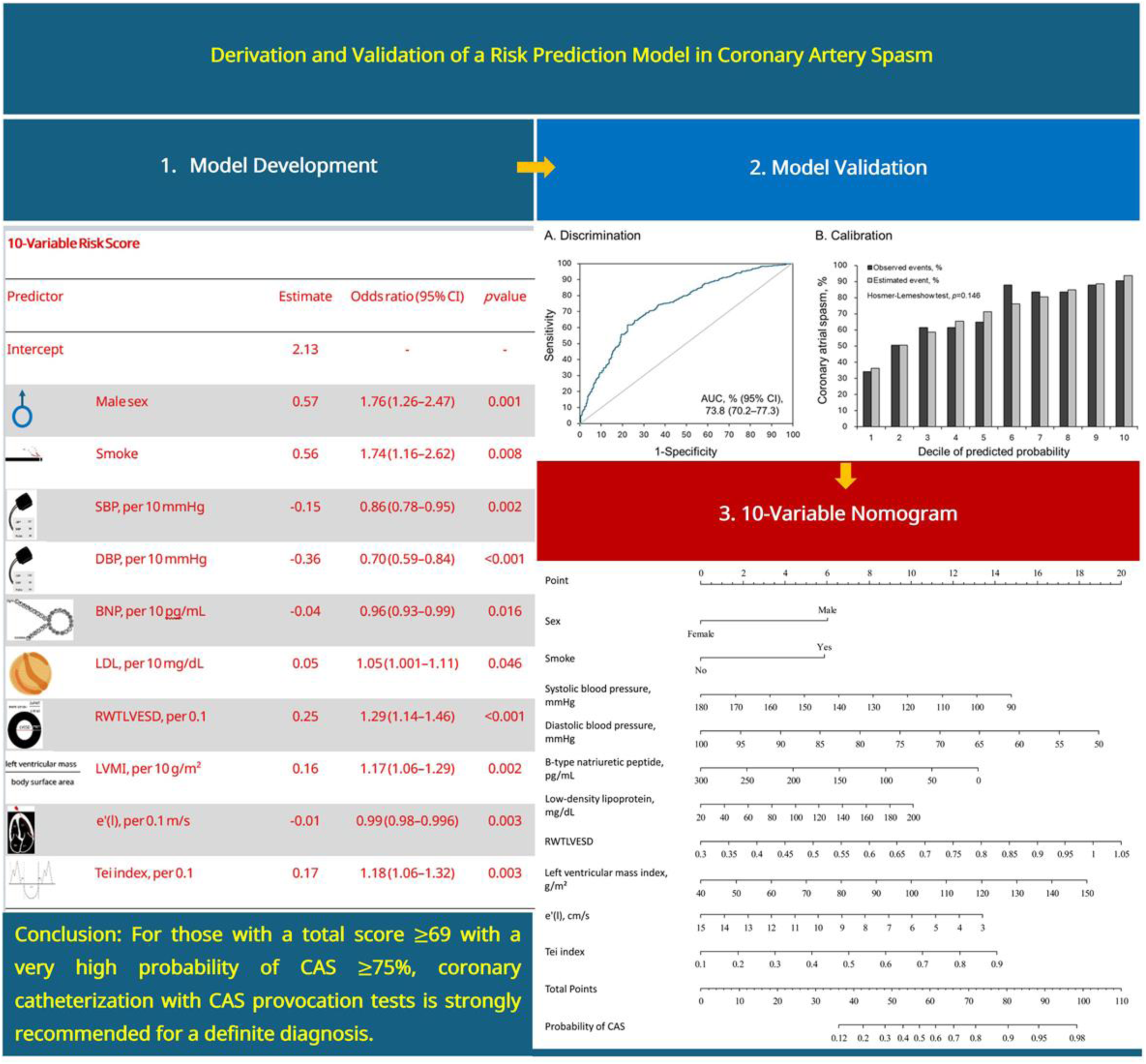

**CLINICAL PERSPECTIVE:** *What Is New?:*

- A simple comprehensive 10-variable scoring model (sex, smoke, systolic and diastolic blood pressure, B-type natriuretic peptide, low-density lipoprotein, left ventricular mass index, relative wall thickness at end-systole, e’(l) and Tei) could offer a non-invasive means of identifying patients at earlier stages of developing coronary artery spasm (CAS).
- While the probability of CAS was <25% when the total score was ≤45, for those with a total score ≥69 with a very high probability of CAS ≥75%, coronary catheterization with CAS provocation tests is strongly recommended for a definite diagnosis.

*What Are the Clinical Implications?:*

- This well-calibrated CAS risk score model has good discrimination, which allows for early allocating diagnostic coronary catheterization effectively, individualized treatments, and better outcomes.

## Introduction

Epicardial large coronary artery spasm (CAS), an extreme vasoconstriction of vascular smooth muscle cell resulting in total or subtotal luminal narrowing, is documented to cause stable or unstable angina, myocardial ischemia or myocardial infarction, and sudden cardiac death.^1,2^ While smoking, age, C-reactive protein (CRP),^3^ aldehyde dehydrogenase 2 deficiency,^4^ and lipoprotein(a)^4^ are risk factors for CAS, CAS is not associated with the classic risk factors such as diabetes mellitus and hypertension^5^ for obstructive coronary artery disease (CAD),^3^ suggesting pathophysiological differences exist between CAS and CAD. The Framingham Heart Study introduced the concept of risk factors in 1961, developed the first formulas to predict cardiovascular complications and algorithms to stratify risk factors in individuals without CAD in 1998, and published other algorithms to estimate global cardiovascular risk in 2008.^6^ These models rely on specific, structured data, such as numeric laboratory values or race to be selected from a multiple choice list, while unstructured data like imaging cannot be incorporated into most of these traditional models; hence, traditional models often oversimplify the complexity of human health, lead to less accurate predictions and can mask the importance of other variables in predicting health outcomes. We have demonstrated that clinicians should aggressively treat CAS patients to ensure timely provision of care due to the high risk of stroke and cardiovascular mortality.^7^ Although invasive CAS provocative testing is the gold standard examination, advances in noninvasive scoring system will be helpful to recognize CAS and guide medical therapy. To date, while only a comprehensive clinical score system with good accuracy in predicting CAS has shown a high area under the curve (AUC) of 0.952,^8^ among which 3 score variables, asthma, ST-segment elevation and the hyperventilation test, require attention. First, less than 4% of CAS patients have asthma.^9^ Second, electrocardiography may appear normal at the beginning of CAS or when the CAS is mild.^3^ Third, CAS is most frequently associated with ST-segment depression rather than elevation.^3^ Fourth, alternating ST-segment elevation and depression could occur in the same patient or even in the same lead within minutes or hours; therefore, the direction and extent of ST-segment deviation may change over time.^3^ Fifth, the hyperventilation provocative test is less sensitive in patients with less frequent CAS attacks and has a danger of inducing simultaneous multi-vessel CAS with this method.^3^ In contrast, over the past 3 decades, several other risk scores for detecting CAD as a measure of prevention have been developed and validated; however, these models often fall short in accounting for the biological complexity and dynamic nature of coronary health, and the lack of a clinically relevant CAS risk prediction model highlights a gap in understanding and treating CAS.

Several cumulative risk factors usually exist together in an individual and interact in a detrimental manner to increase a person’s chance of getting CAS,^2^ suggesting a multifactorial approach considering all the risk factors may be the best strategy for the prevention of CAS. Before referral for the gold standard diagnostic method of coronary angiography with provocative testing, estimating CAS risk often relies on a set of factors including age, sex, medical history, smoking exposure, CRP levels, symptoms such as chest pain occurring at rest or from overnight into early morning, and ischemic electrocardiographic changes during attacks.^3^ In addition, because B-type natriuretic peptide (BNP), secreted by ventricular cardiomyocytes, has potent vasodilatory effects predominantly on epicardial conductance vessels, it raises the possibility of its deficiency in patients with epicardial CAS.^10^ Gohbara et al. have demonstrated that BNP is lower in CAS-induced than CAD-induced non-progressive ST-elevation acute coronary syndrome.^11^ Therefore, we hypothesized that BNP could differentiate CAS-related from non-CAS-related angina in patients without obstructive CAD. On the other hand, abnormal left ventricular (LV) filling and relaxation are earliest detectable sensitive signs of myocardial ischemia, which are reversible after interventions to restore coronary flow.^12^ Hence, it is reasonable to assume that good quantitative echocardiographic analysis should identify significant differences between CAS-induced and non-CAS-induced angina. Notably, in the assessment of global systolic and diastolic function, the Tei index is a simple doppler parameter independent of heart rate and blood pressure, applies to left and right ventricular systolic and diastolic dysfunction, does not rely on geometric assumptions, and is highly reproducible.^13^ As the endothelium-dependent vasodilation is impaired progressively with abnormal LV geometry,^14^ and non-inflamed posterior wall has been shown to undergo hypertrophy during anterior wall myocardial infarction in mice, remote tissue can be adversely affected by inflammation from the site of ischemia.^15^ Furthermore, relative wall thickness (RWT), the reference value from posterior wall thickness, is a cardiovascular risk factor.^16^ LV geometric patterns based on RWT are also believed to have incremental value for predicting cardiovascular events.^17^ Little is known, however, about whether patients with CAS are more likely to have abnormal remodeling or LV relaxation.

Accurate prediction and early diagnosis of CAS are crucial for managing and preventing cardiovascular events, especially in individuals with multiple mild abnormalities. In CAS, it is uncommon for subjects to have more than 4 risk factors, and estimates of CAS risk tend to be more precise for individuals with fewer risk factors. Moreover, no clinical diagnostic score incorporating echocardiographic parameters has been investigated for CAS. While predicting CAS in a free-living population not on medication is emphasized, we aimed to evaluate the utility and accuracy of a risk score including clinical characteristics, blood biochemistry and echocardiographic parameters to identify undiagnosed CAS but without obstructive CAD in primary care.

## Methods

### Study Population

From November 2008 to March 2025, 913 Taiwanese patients with suspected ischemic heart disease but without angiographic evidence of obstructive CAD were subjected to intracoronary methylergonovine testing in Shuang Ho Hospital, Taipei Medical University. Patients were grouped according to the presence or absence of CAS. Diagnostic criteria for CAS included spontaneous chest pain at rest, ST-segment elevation or depression on electrocardiogram that was relieved by sublingual administration of nitroglycerin, and a positive result on intracoronary methylergonovine provocation testing. The control group consisted of patients who presented with atypical chest pain which was not provoked by exertion and had negative results on intracoronary methylergonovine provocation testing. Exclusion criteria included obstructive CAD, previous coronary angioplasty or myocardial infarction, coronary microvascular spasm,^18^ severe valvular heart disease, inflammatory manifestations probably associated with noncardiac diseases (e.g., infections and autoimmune disorders), liver disease/renal failure (serum creatinine level >2.5 mg/dL), collagen disease, malignancy and missing blood samples. None of our patients had allergic or hypersensitivity conditions. This study was approved by the Taipei Medical University-Joint Institutional Review Board (approval number: 201011004) and all patients gave written informed consent.

### Clinical Data

Patients were assessed for the presence of the following cardiac risk factors: age, sex, cigarette smoking status, diabetes mellitus, hypercholesterolemia, and hypertension. Current smoking status was defined as at least 0.5 pack year and having smoked at least 1 cigarette within 3 weeks of cardiac catheterization. Patients were determined to have diabetes mellitus if they were currently on dietary treatment and/or medical therapy for diabetes mellitus. The average baseline self-measured systolic and diastolic seated blood pressure at home was the mean of a minimum 12 readings total optimally for 7 days with a minimum of 3 days.^19^ Hypertension was defined as blood pressure of >130/80 mm Hg or receiving antihypertensive treatment. Hypercholesterolemia was diagnosed in patients with serum total cholesterol >200 mg/dL. All patients underwent echocardiography, when heart rates were recorded, before coronary angiography and within 2 weeks of the last angina. The blood pressure levels were measured at the time of coronary angiography.

### Laboratory Analysis

Data for serum creatinine, estimated glomerular filtration rate, hemoglobin, hematocrit, platelet counts, white blood cell count, monocyte counts, BNP, high sensitivity C-reactive protein (hs-CRP), blood glucose, hemoglobin A1c, total cholesterol, triglycerides, high-density lipoprotein and low-density lipoprotein were obtained on admission.

### Coronary Angiography and Intracoronary Methylergonovine Testing

Coronary angiography was performed within 2 months of chest pain using the standard Judkins technique via a femoral or a radial approach. Nitrates and calcium antagonists were withdrawn for ≥24 hours before coronary angiography. The LV ejection fraction was calculated using Simpson’s method. Selective left and right coronary angiography were performed in multiple axial and hemiaxial projections. Obstructive CAD was defined as ≥50% diameter reduction in lumen caliber after administration of intracoronary nitroglycerin (100 μg).^20^ Intracoronary methylergonovine (Methergin®; Novartis, Basel, Switzerland) provocation testing was performed in succession if no obstructive CAD was found. Methylergonovine was administered stepwise (1, 5, 10, 30 μg) first into the right coronary artery and subsequently into the left coronary artery. Provocation testing for CAS was considered positive when there was a >70% reduction in luminal diameter compared to post intracoronary nitroglycerin and when there was associated angina and/or ST depression or elevation.^21^ Provocation testing was stopped with intracoronary nitroglycerin 100-200 μg (Millisrol®; G. Pohl-Boskamp, Hohenlockstedt, Germany). The observation of reversal changes in the coronary artery diameter further confirmed the diagnosis of CAS. Spontaneous CAS was defined as the relief of >70% diameter stenosis after intracoronary nitroglycerin 100-200 μg administration.

### Echocardiographic Methods

Echocardiograms were obtained by using a Philips iE33 (Philips Ultrasound, Bothell, Washington, USA), with the patients breathing quietly in the left decubitus position. The patients underwent 2-dimensional and M-mode echocardiographic examinations. left atrial antero-posterior and LV diameters, wall thickness and ejection fraction were measured according to the guidelines of the American Society of Echocardiography.^22,23^ The LV ejection fraction^23^ was calculated according to modified Simpson’s rules.^24^ End-diastolic LV dimensions were used to calculate LV mass by an anatomically validated formula.^25^ Left atrial (LA) diameter was measured in the parasternal long-axis view from the trailing edge of the posterior aortic-anterior LA complex at end systole as the anteroposterior linear diameter, using 2-dimensional echocardiographic guidance to position the cursor as recommended.^26^ RWT at end-diastole (RWTLVEDD) was calculated as posterior wall thickness/internal radius.^27^ Increased RWTLVEDD was regarded present when the ratio was ≥0.43.^28^ RWT at end-systole (RWTLVESD) was also calculated by a similar formula because this contributes to the LV geometric component of passive ventricular stiffness that is superimposed on the active relaxation in early diastole.^16^ LV hypertrophy was considered present when LV mass indexed (LVMI) for body surface area was >116 g/m^2^ for men and >104 g/m^2^ for women.^29^ The combinations of LVMI and RWTLVEDD defined 4 LV geometric patterns: normal geometry, concentric hypertrophy with increased RWTLVEDD and LV hypertrophy, eccentric hypertrophy with normal RWTLVEDD and LV hypertrophy, and concentric remodeling with increased RWTLVEDD and normal LVMI.^30^

Peak velocities of early diastolic filling (E), late diastolic filling (A), and deceleration time were derived from transmitral Doppler recordings. Isovolumic relaxation time (IVRT) was measured at the apical 5-chamber view with the sampling volume positioned between the mitral valve and the LV outflow tract as the time taken from the closure of the aortic valve to the opening of mitral valve. Doppler tissue echocardiography was performed at transducer frequencies of 3.5–4.0 MHz, adjusting the spectral pulsed Doppler signal filters until a Nyquist limit of 15–20 cm/s was reached, and using the minimum optimal gain. The monitor sweep speed was set at 50–100 mm/s to optimize the spectral display of myocardial velocities. Tissue Doppler-derived early diastolic velocities (e’), and late diastolic velocities (a’) were derived from the lateral and medial mitral annulus. The lateral and mitral E/e’ ratio was subsequently calculated. While τ, the time constant of the LV pressure decay at isovolumic relaxation, is a sensitive index of myocardial ischemia, τ with a zero asymptote assumption (τ_0_) was estimated using the following equation: τ_0_(ms) = IVRT_doppler_/[ln(P_s_)-ln(P_PCWP_)],^31^ where P_s_ is systolic blood pressure and PCWP is mean pulmonary capillary wedge pressure. All procedures complied with the guidelines from the American Society of Echocardiography.^32^

### Statistical Analysis

Baseline characteristics between the CAS and control groups were compared using the chi-square test for categorical variables, the independent sample t-test for continuous variables, and the Mann-Whitney U-test for BNP and hs-CRP due to their non-normal distribution. The potential predictive factors comprised 52 parameters, including 8 demographic variables (e.g., age, sex, smoking status), 18 vital signs and laboratory results (e.g., systolic blood pressure, creatinine), and 26 echocardiographic features, as detailed in **Table 1**. In the first step, univariate logistic regression analyses were conducted to identify potential factors associated with CAS risk based on all baseline characteristics. Variables with a significance level of less than 0.15 in the univariate analyses were then included in a multivariable logistic regression model with backward elimination. Discriminative performance was evaluated using the area under the curve (AUC), with an AUC value above 0.7 considered acceptable.^33^

**Table 1.** Baseline Characteristics of Patients in the CAS and non-CAS Control Groups.

| Variable | Total<br>(*n* = 913) | CAS<br>(*n* = 645) | Control<br>(*n* = 268) | *p* value |
| --- | --- | --- | --- | --- |
| Age, year | 56.4 ± 12.6 | 57.1 ± 11.9 | 54.8 ± 14.2 | 0.014 |
| Male | 453 (49.6) | 352 (54.6) | 101 (37.7) | <0.001 |
| Body mass index, kg/m<sup>2</sup> | 26.1 ± 4.4 | 26.2 ± 4.2 | 25.8 ± 4.6 | 0.187 |
| Weight status |  |  |  | 0.097 |
| Normal/Lean (<23) | 226 (24.8) | 147 (22.8) | 79 (29.5) |  |
| Overweight (23-24.9) | 174 (19.1) | 124 (19.2) | 50 (18.7) |  |
| Obese (≥25) | 513 (56.2) | 374 (58.0) | 139 (51.9) |  |
| Body surface area, m<sup>2</sup> | 1.76 ± 0.20 | 1.77 ± 0.20 | 1.73 ± 0.21 | 0.009 |
| Smoke | 229 (25.1) | 184 (28.5) | 45 (16.8) | <0.001 |
| Diabetes mellitus | 109 (11.9) | 81 (12.6) | 28 (10.4) | 0.433 |
| Hypertension | 301 (33.0) | 220 (34.1) | 81 (30.2) | 0.279 |
| Systolic blood pressure, mmHg | 122.0 ± 18.9 | 120.2 ± 17.7 | 126.3 ± 20.9 | <0.001 |
| Diastolic blood pressure, mmHg | 74.3 ± 10.6 | 73.4 ± 10.3 | 76.6 ± 11.0 | <0.001 |
| Heart rate, bpm | 69.1 ± 11.5 | 68.7 ± 11.7 | 70.2 ± 11.1 | 0.072 |
| Serum creatinine, mg/dL | 0.86 ± 0.31 | 0.85 ± 0.27 | 0.87 ± 0.39 | 0.520 |
| Estimated glomerular filtration rate, mL/min/1.73 m<sup>2</sup> | 91.6 ± 23.1 | 92.2 ± 22.0 | 90.1 ± 25.6 | 0.227 |
| Hemoglobin, g/dL | 13.6 ± 1.5 | 13.8 ± 1.5 | 13.3 ± 1.5 | <0.001 |
| Hematocrit, % | 40.1 ± 4.4 | 40.6 ± 4.2 | 39.0 ± 4.5 | <0.001 |
| Platelet counts, 10<sup>9</sup>/L | 231.4 ± 58.7 | 232.0 ± 59.4 | 229.9 ± 57.1 | 0.618 |
| White blood cell count, 10<sup>9</sup>/L | 6822.6 ± 1730.9 | 6874.0 ± 1790.0 | 6698.8 ± 1576.1 | 0.164 |
| Monocyte counts, 10<sup>3</sup>/L | 495.2 ± 165.2 | 501.9 ± 168.4 | 479.2 ± 156.5 | 0.058 |
| B-type natriuretic peptide, pg/mL |  |  |  |  |
| Mean ± standard deviation | 35.8 ± 49.5 | 33.6 ± 45.3 | 41.0 ± 58.1 | 0.038 |
| Median [25th, 75th percentile] | 20.9 [9.0, 42.8] | 20.3 [7.6, 40.0] | 23.0 [10.3, 48.5] | 0.078 |
| High-sensitivity C-reactive protein, mg/L |  |  |  |  |

| Variable | Total<br>(n = 913) | CAS<br>(n = 645) | Control<br>(n = 268) | p value |
| --- | --- | --- | --- | --- |
| Mean ± standard deviation | 2.03 ± 3.62 | 2.14 ± 3.76 | 1.75 ± 3.26 | 0.135 |
| Median [25th, 75th percentile] | 0.82 [0.40, 1.91] | 0.90 [0.40, 2.01] | 0.70 [0.39, 1.64] | 0.074 |
| High-sensitivity C-reactive protein, mg/L |  |  |  | 0.112 |
| ≤1 | 505 (55.3) | 344 (53.3) | 161 (60.1) |  |
| 1-2.9 | 266 (29.1) | 192 (29.8) | 74 (27.6) |  |
| ≥3 | 142 (15.6) | 109 (16.9) | 33 (12.3) |  |
| Fasting glucose, mg/dL | 100.3 ± 19.5 | 101.0 ± 19.7 | 98.8 ± 18.8 | 0.118 |
| Glycated hemoglobin, % | 5.76 ± 0.66 | 5.79 ± 0.68 | 5.71 ± 0.61 | 0.112 |
| Total cholesterol, mg/dL | 170.6 ± 36.4 | 171.7 ± 36.5 | 168.0 ± 35.9 | 0.160 |
| Triglycerides, mg/dL | 122.4 ± 83.9 | 126.8 ± 86.9 | 112.0 ± 75.3 | 0.015 |
| High-density lipoprotein, mg/dL | 46.2 ± 12.8 | 45.6 ± 12.4 | 47.6 ± 13.7 | 0.033 |
| Low-density lipoprotein, mg/dL | 100.4 ± 31.2 | 101.6 ± 31.0 | 97.6 ± 31.4 | 0.075 |
| Echocardiography |  |  |  |  |

|  | Total | CAS | Control |  |
| --- | --- | --- | --- | --- |
| Variable | (n = 913) | (n = 645) | (n = 268) | p value |
| LV ejection fraction, % | 65.4 ± 6.9 | 65.6 ± 6.8 | 64.9 ± 7.3 | 0.154 |
| Fractional shortening | 0.36 ± 0.06 | 0.36 ± 0.06 | 0.36 ± 0.06 | 0.403 |
| Left atrium diameter, mm | 38.6 ± 6.1 | 38.8 ± 6.0 | 38.2 ± 6.2 | 0.159 |
| Interventricular septal thickness, mm | 9.18 ± 1.81 | 9.28 ± 1.74 | 8.95 ± 1.96 | 0.014 |
| LVEDD, mm | 44.4 ± 4.7 | 44.6 ± 4.6 | 44.1 ± 4.7 | 0.139 |
| LVESD, mm | 28.4 ± 4.2 | 28.4 ± 4.2 | 28.2 ± 4.1 | 0.406 |
| Left ventricular posterior wall, mm | 8.41 ± 1.55 | 8.63 ± 1.44 | 7.89 ± 1.66 | <0.001 |
| RWTLVEDD | 0.38 ± 0.08 | 0.39 ± 0.08 | 0.36 ± 0.08 | <0.001 |
| RWTLVESD | 0.60 ± 0.14 | 0.62 ± 0.13 | 0.57 ± 0.14 | <0.001 |
| LV mass, g | 131.3 ± 39.5 | 134.8 ± 37.9 | 122.8 ± 42.0 | <0.001 |
| LV mass index, g/m<sup>2</sup> | 74.3 ± 19.4 | 76.0 ± 18.6 | 70.4 ± 20.9 | <0.001 |
| LV remodeling |  |  |  | 0.012 |
| Normal | 633 (69.3) | 429 (66.5) | 204 (76.1) |  |

| Variable | Total<br>(n = 913) | CAS<br>(n = 645) | Control<br>(n = 268) | p value |
| --- | --- | --- | --- | --- |
| Concentric remodeling | 239 (26.2) | 187 (29.0) | 52 (19.4) |  |
| Concentric hypertrophy | 26 (2.8) | 20 (3.1) | 6 (2.2) |  |
| Eccentric hypertrophy | 15 (1.6) | 9 (1.4) | 6 (2.2) |  |
| Mitral inflow deceleration time, ms | 188.7 ± 34.6 | 189.9 ± 34.5 | 185.8 ± 34.9 | 0.100 |
| Isovolumic relaxation time, ms | 86.5 ± 12.6 | 87.3 ± 12.5 | 84.6 ± 12.7 | 0.003 |
| $\tau_{0l}$: LV relaxation time constant using lateral annulus, ms | 37.8 ± 6.8 | 38.4 ± 6.6 | 36.4 ± 7.1 | <0.001 |
| $\tau_{0m}$: LV relaxation time constant using medial annulus, ms | 40.4 ± 7.6 | 40.9 ± 7.3 | 39.3 ± 8.1 | 0.004 |
| E: Mitral inflow E-wave velocity, cm/s | 60.8 ± 15.5 | 59.5 ± 15.2 | 64.0 ± 15.7 | <0.001 |
| A: Mitral inflow A-wave velocity, cm/s | 59.1 ± 16.9 | 59.0 ± 16.2 | 59.3 ± 18.5 | 0.828 |
| E/A: Mitral inflow E/A ratio | 1.11 ± 0.44 | 1.08 ± 0.41 | 1.19 ± 0.51 | 0.001 |
| e'(l): Lateral mitral annular early diastolic velocity, cm/s | 7.95 ± 2.54 | 7.73 ± 2.41 | 8.47 ± 2.77 | <0.001 |
| a'(l): Lateral mitral annular late diastolic velocity, cm/s | 9.36 ± 2.26 | 9.28 ± 2.21 | 9.55 ± 2.37 | 0.103 |
| e'(m): Medial mitral annular early diastolic velocity, cm/s | 6.38 ± 1.93 | 6.27 ± 1.86 | 6.63 ± 2.08 | 0.010 |

|  | Total | CAS | Control |  |
| --- | --- | --- | --- | --- |
| Variable | (*n* = 913) | (*n* = 645) | (*n* = 268) | *p* value |
| a'(m): Medial mitral annular late diastolic velocity, cm/s | 8.76 ± 1.72 | 8.76 ± 1.69 | 8.74 ± 1.77 | 0.863 |
| E/e'(l): Lateral mitral inflow E to e' ratio | 8.25 ± 2.95 | 8.27 ± 2.89 | 8.20 ± 3.07 | 0.760 |
| E/e'(m): Medial mitral inflow E to e' ratio | 10.20 ± 3.46 | 10.09 ± 3.28 | 10.46 ± 3.87 | 0.146 |
| Tei index | 0.48 ± 0.16 | 0.50 ± 0.15 | 0.45 ± 0.15 | <0.001 |

Data are presented as frequency (percentage), mean ± standard deviation or median [25<sup>th</sup> percentile, 75<sup>th</sup> percentile].
CAS indicates coronary artery spasm; LV, left ventricle; LVEDD, left ventricular end-diastolic diameter; LVESD, left ventricular end-systolic diameter;
RWTLVEDD, relative wall thickness using LVEDD; and RWTLVESD, relative wall thickness using LVESD.

The performance of calibration was evaluated by comparing the predicted probability to the actual observed probability across the deciles of the predicted probabilities. Calibration performance was assessed by comparing predicted probabilities to actual observed probabilities across deciles of the predicted values, supplemented by the Hosmer-Lemeshow goodness-of-fit test. The generalizability of the obtained model was assessed through internal validation using 1,000 bootstrap samples, providing reasonably valid estimates of expected optimism.^34^ At last, the results of the final multivariable logistic regression model were converted into a simplified point system to enhance clinical applicability. The key concept involves rounding off regression coefficients, with further details available in a previous report.^35^ All tests were 2-tailed and *P* <0.05 was considered statistically significant. Data analyses were conducted using R, version 4.2.2 (R Foundation for Statistical Computing) with package ‘rms’ (Frank E. Harrell Jr).

## Results

### Patient Characteristics

A total of 913 patients with complete baseline data were included in the study, comprising 645 (70.6%) CAS cases and 268 (29.4%) non-CAS controls. Table 1 presents detailed baseline characteristic data. The mean age was 57.1 years in the CAS group and 54.8 years in the control group, with a significant difference (p=0.014). The proportion of male participants was higher in the CAS group (54.6% vs. 37.7%, p<0.001). Regarding other demographics, vital signs, and laboratory results, patients with CAS had a larger body surface area, were more likely to smoke, had lower systolic and diastolic blood pressure levels, had higher hemoglobin and hematocrit levels, lower B-type natriuretic peptide (BNP) levels, elevated triglycerides and low-density lipoprotein (LDL) levels, and lower high-density lipoprotein levels (p<0.05). Several echocardiography parameters differed significantly between groups, as detailed in Table 1.

### Associated Factors of CAS

Table 2 presents the univariate logistic regression analyses for the preliminary screening of potential factors associated with CAS. The final multivariable logistic regression model identified male sex, smoking, lower systolic and diastolic blood pressure, lower BNP levels, higher LDL levels, greater RWTLVESD, larger LVMI, lower e’(l) values and a higher Tei index as significant factors associated with CAS (p<0.05; Table 3). The performance of discrimination was modest with the AUC value of 73.8% (Figure 1A). After correcting for optimism through bootstrapped internal validation, the AUC slightly decreased to 72.4%, suggesting the potential generalizability of the derived model (Data not shown). The model also demonstrated good calibration, with non-substantial discrepancy (p of Hosmer-Lemeshow test=0.146) between predicted and actual probabilities (Figure 1B).

**Figure 1.**
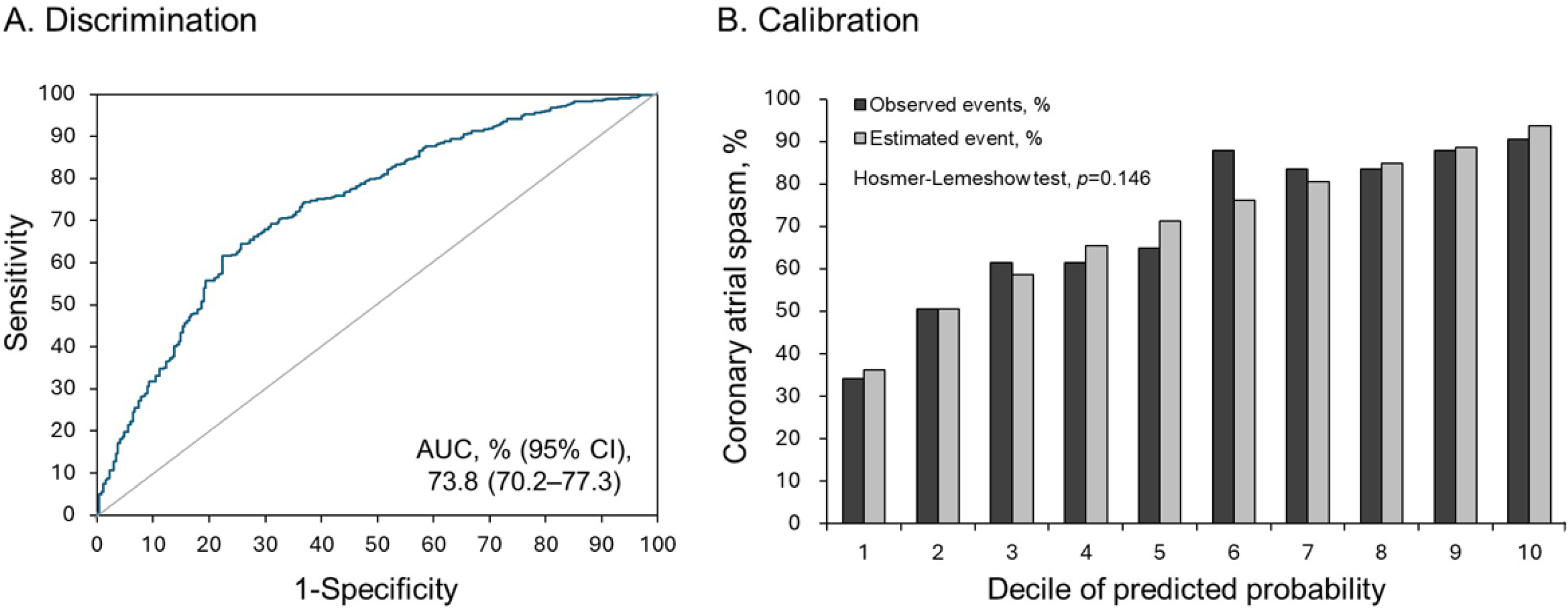
Discrimination (A) and calibration (B) performance of the derived multivariable logistic regression model for predicting CAS. AUC, area under the curve; CAS, coronary artery spasm; CI, confidence interval.

**Table 2.**
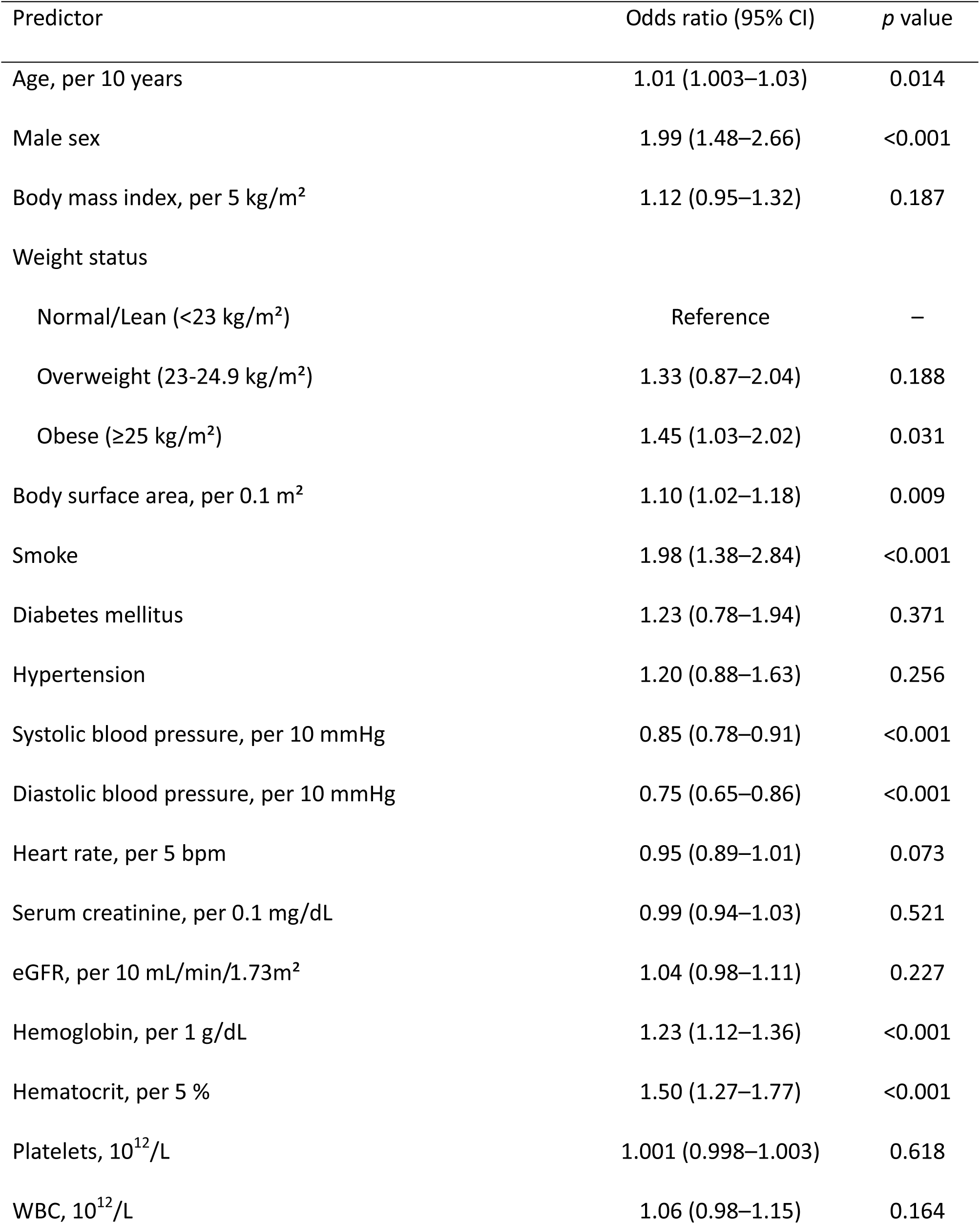
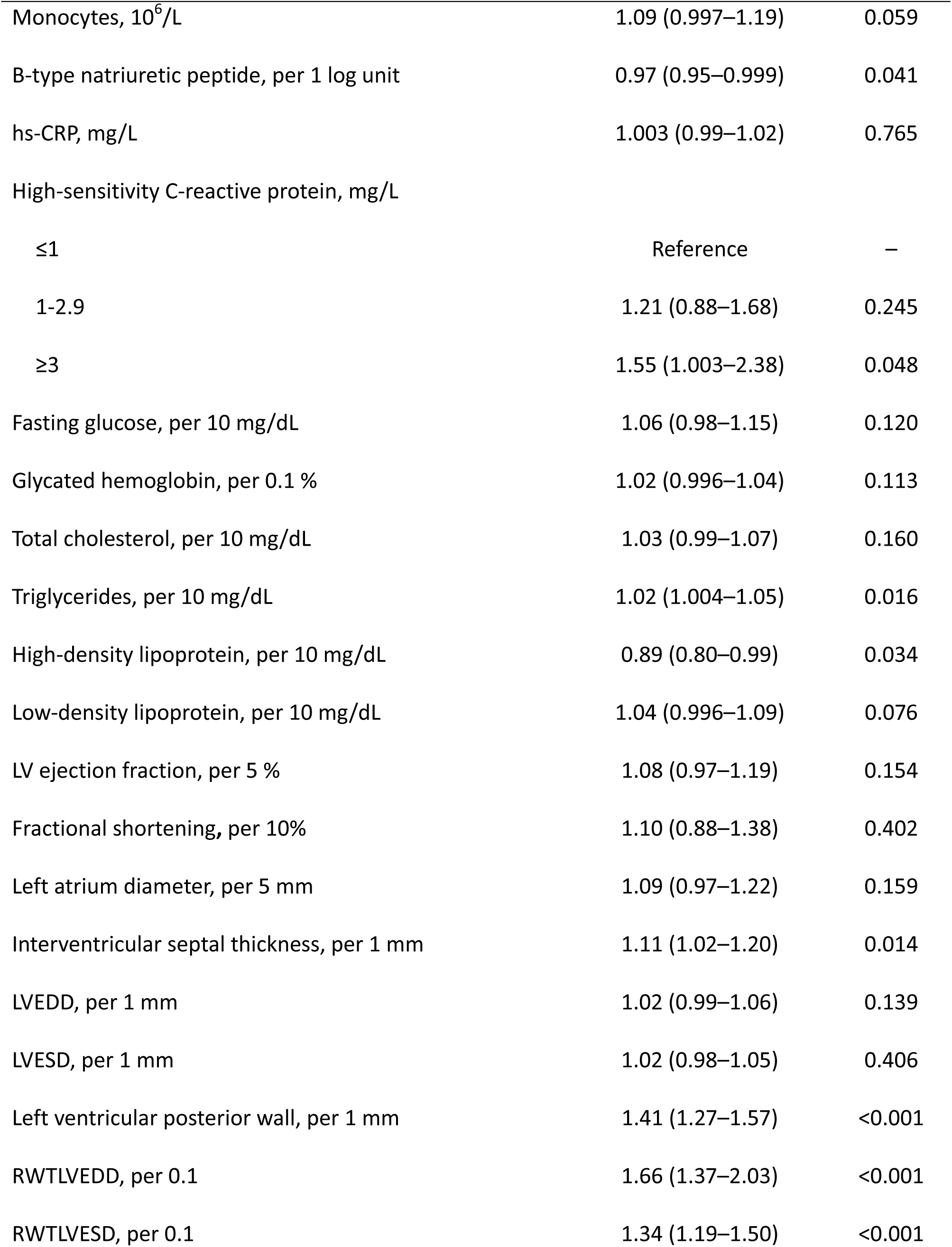
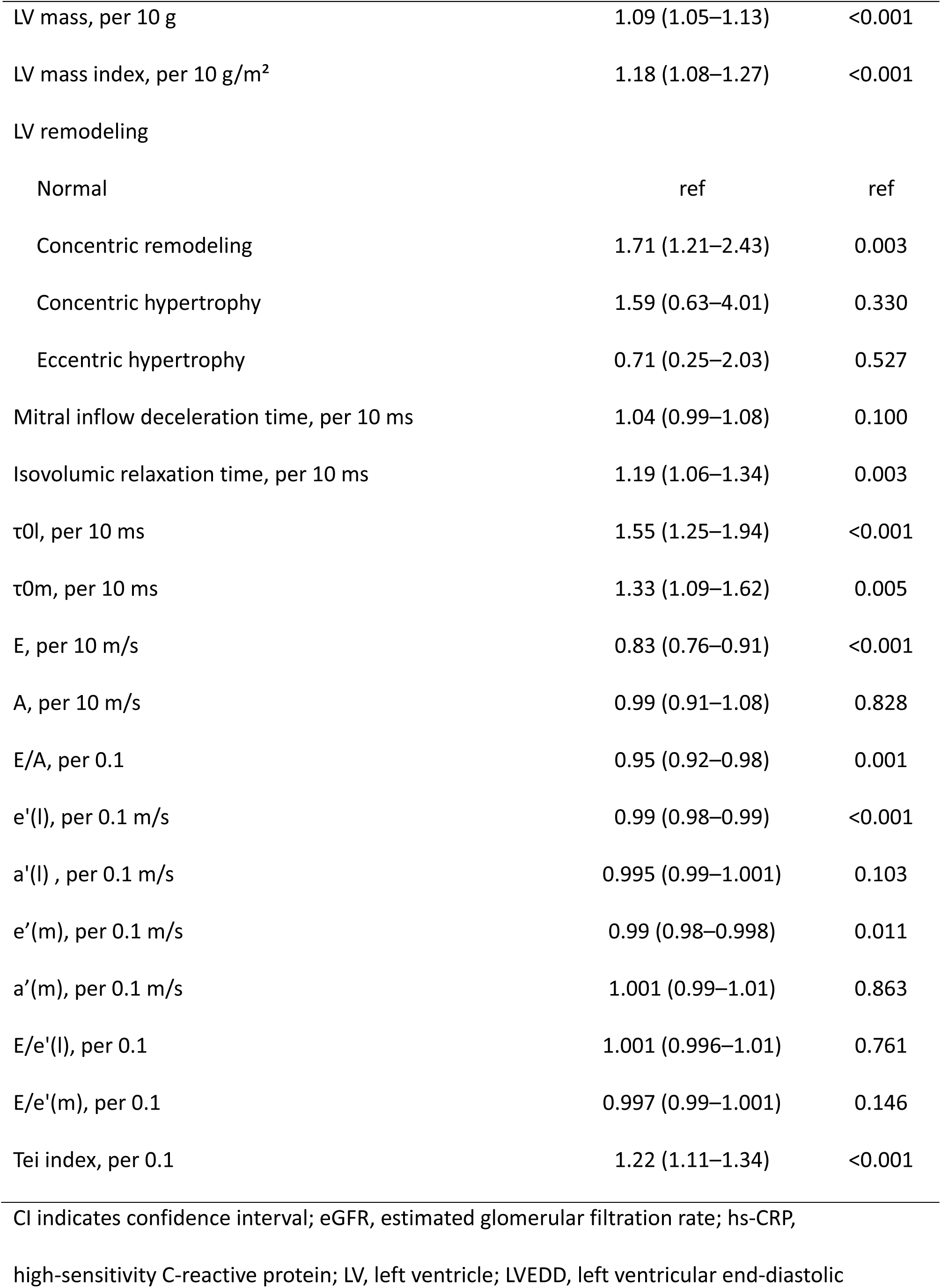
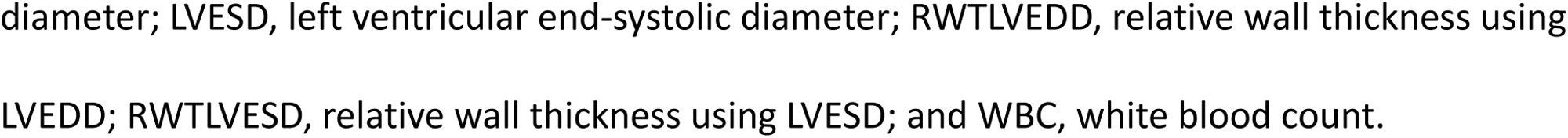
Univariate Logistic Regression Analysis of the Potential Association Between the Baseline Characteristics and Risk of Coronary Artery Spasm.

**Table 3.**
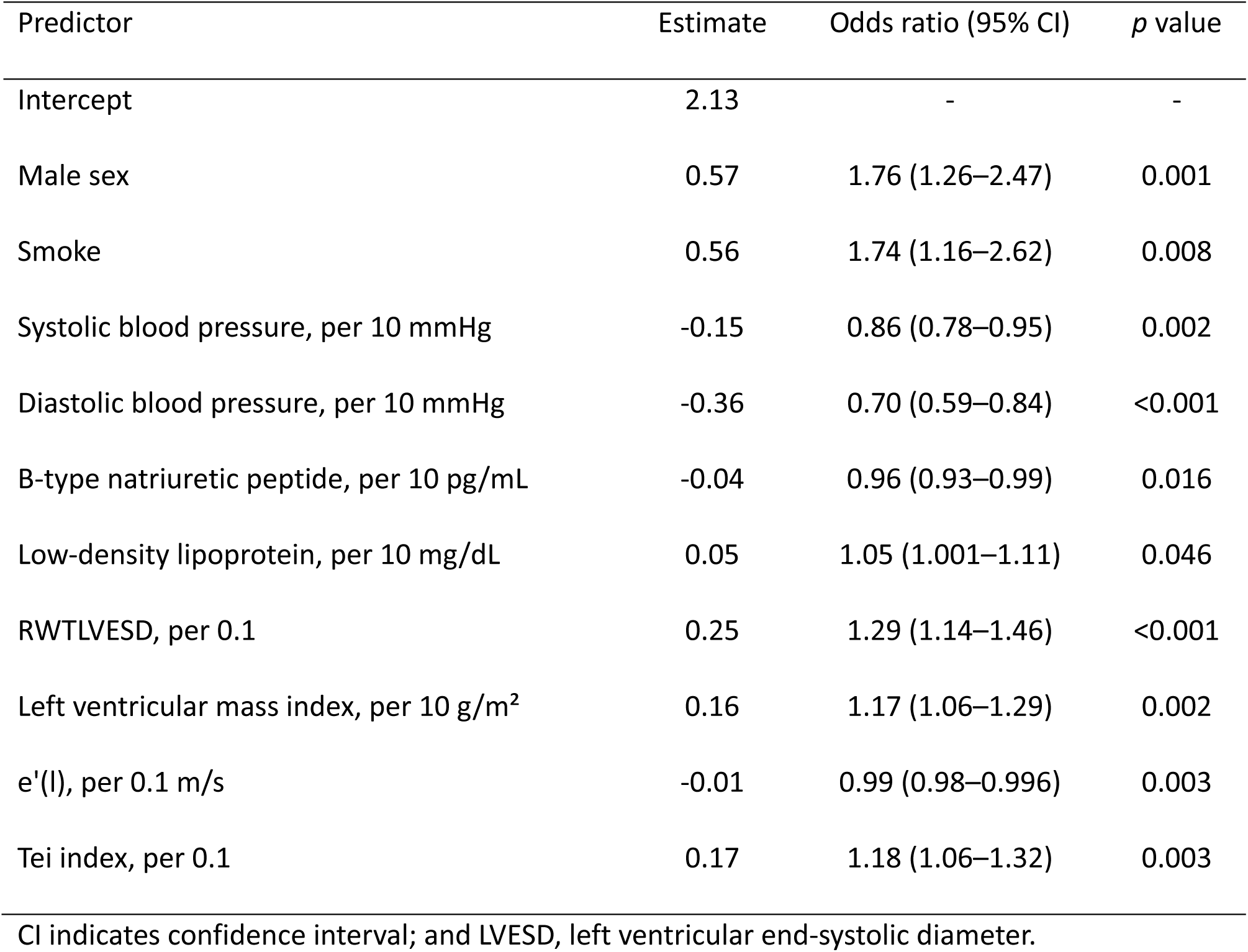
Multivariable logistic regression analysis of the associated factors with risk of coronary artery spasm.

| Predictor | Estimate | Odds ratio (95% CI) | *p* value |
| --- | --- | --- | --- |
| Intercept | 2.13 | - | - |
| Male sex | 0.57 | 1.76 (1.26–2.47) | 0.001 |
| Smoke | 0.56 | 1.74 (1.16–2.62) | 0.008 |
| Systolic blood pressure, per 10 mmHg | -0.15 | 0.86 (0.78–0.95) | 0.002 |
| Diastolic blood pressure, per 10 mmHg | -0.36 | 0.70 (0.59–0.84) | <0.001 |
| B-type natriuretic peptide, per 10 pg/mL | -0.04 | 0.96 (0.93–0.99) | 0.016 |
| Low-density lipoprotein, per 10 mg/dL | 0.05 | 1.05 (1.001–1.11) | 0.046 |
| RWTLVESD, per 0.1 | 0.25 | 1.29 (1.14–1.46) | <0.001 |
| Left ventricular mass index, per 10 g/m<sup>2</sup> | 0.16 | 1.17 (1.06–1.29) | 0.002 |
| e'(l), per 0.1 m/s | -0.01 | 0.99 (0.98–0.996) | 0.003 |
| Tei index, per 0.1 | 0.17 | 1.18 (1.06–1.32) | 0.003 |

### Clinical Use of the Prediction Model

Table 4 presents the simplified scoring function for CAS prediction, derived from the multivariable logistic regression model. The total score ranged from 36 to 98, corresponding to predicted CAS probabilities of 12% and 98%, respectively. Although a simplified scoring function offers ease of manual calculation, a nomogram provides a more flexible and visually intuitive tool that incorporates continuous variables and nonlinear relationships, enhancing individualized risk assessment and clinical decision-making (Figure 2).

**Figure 2.**
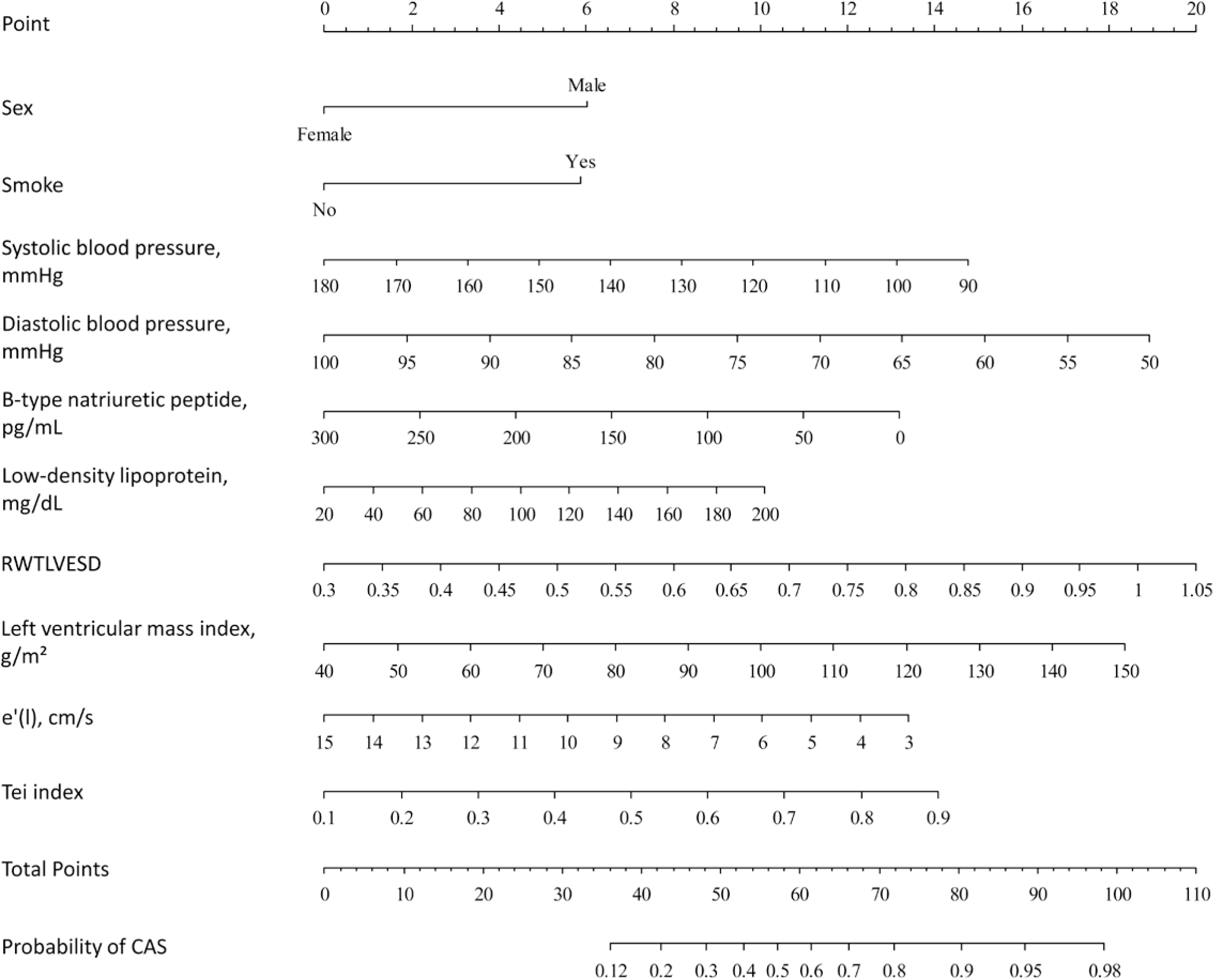
Nomogram illustrating the relationships between predictive variables and the probability of CAS, derived from the multivariable logistic regression model. CAS, coronary artery spasm; LVESD, left ventricular end-systolic diameter.

**Table 4.** Simplified score function of the prediction model for coronary artery spasm.

| Point values for each variable |  |  |  | Summation of points |  |
| --- | --- | --- | --- | --- | --- |
| Variable | Point | Variable | Point | Total points | Probability of CAS |
| **Sex** |  | **RWTLVESD** |  | 36 | 0.12 |
| Female | 0 | <0.35 | 0 | 37 | 0.13 |
| Male | 6 | 0.35-0.39 | 1 | 38 | 0.14 |
| **Smoke** |  | 0.40-0.44 | 3 | 39 | 0.15 |
| No | 0 | 0.45-0.49 | 4 | 40 | 0.16 |
| Yes | 6 | 0.50-0.54 | 5 | 41 | 0.18 |
| **SBP, mmHg** |  | 0.55-0.59 | 7 | 42 | 0.19 |
| <100 | 15 | 0.60-0.64 | 8 | 43 | 0.21 |
| 100-109 | 13 | 0.65-0.69 | 9 | 44 | 0.22 |
| 110-119 | 11 | 0.70-0.74 | 11 | 45 | 0.24 |
| 120-129 | 10 | 0.75-0.79 | 12 | 46 | 0.26 |
| 130-139 | 8 | 0.80-0.84 | 13 | 47 | 0.27 |
| 140-149 | 7 | 0.85-0.89 | 15 | 48 | 0.29 |
| 150-159 | 5 | 0.90-0.94 | 16 | 49 | 0.31 |
| 160-169 | 3 | 0.95-0.99 | 17 | 50 | 0.33 |
| 170-179 | 2 | 1-1.04 | 19 | 51 | 0.35 |
| ≥180 | 0 | ≥1.05 | 20 | 52 | 0.38 |
| **DBP, mmHg** |  | **LVMI, g/m<sup>2</sup>** |  | 53 | 0.40 |
| <55 | 19 | <50 | 0 | 54 | 0.42 |
|  |  |  |  | Total | Probability of |
| Variable | Point | Variable | Point | points | CAS |
| 55-59 | 17 | 50-59 | 2 | 55 | 0.44 |
| 60-64 | 15 | 60-69 | 3 | 56 | 0.47 |
| 65-69 | 13 | 70-79 | 5 | 57 | 0.49 |
| 70-74 | 11 | 80-89 | 7 | 58 | 0.51 |
| 75-79 | 9 | 90-99 | 8 | 59 | 0.54 |
| 80-84 | 8 | 100-109 | 10 | 60 | 0.56 |
| 85-89 | 6 | 110-119 | 12 | 61 | 0.58 |
| 90-94 | 4 | 120-129 | 13 | 62 | 0.61 |
| 95-99 | 2 | 130-139 | 15 | 63 | 0.63 |
| ≥100 | 0 | 140-149 | 17 | 64 | 0.65 |
| **BNP, pg/mL** |  | ≥150 | 18 | 65 | 0.67 |
| <50 | 13 | **e'(l), cm/s** |  | 66 | 0.69 |
| 50-99 | 11 | 3 | 13 | 67 | 0.71 |
| 100-149 | 9 | 4 | 12 | 68 | 0.73 |
| 150-199 | 7 | 5 | 11 | 69 | 0.75 |
| 200-249 | 4 | 6 | 10 | 70 | 0.77 |
| 250-299 | 2 | 7 | 9 | 71 | 0.78 |
| ≥300 | 0 | 8 | 8 | 72 | 0.80 |
| **LDL, mg/dL** |  | 9 | 7 | 73 | 0.81 |
| <30 | 0 | 10 | 6 | 74 | 0.83 |
|  |  |  |  | Total | Probability of |
| Variable | Point | Variable | Point | points | CAS |
| 31-40 | 1 | 11 | 4 | 75 | 0.84 |
| 41-60 | 2 | 12 | 3 | 76 | 0.85 |
| 61-80 | 3 | 13 | 2 | 77 | 0.87 |
| 81-100 | 4 | 14 | 1 | 78 | 0.88 |
| 101-120 | 6 | 15 | 0 | 79 | 0.89 |
| 121-140 | 7 | **Tei index** |  | 80 | 0.90 |
| 141-160 | 8 | <0.2 | 0 | 82 | 0.91 |
| 161-180 | 9 | 0.2 | 2 | 83 | 0.92 |
| ≥180 | 10 | 0.3 | 4 | 85 | 0.93 |
|  |  | 0.4 | 5 | 86 | 0.94 |
|  |  | 0.5 | 7 | 88 | 0.95 |
|  |  | 0.6 | 9 | 91 | 0.96 |
|  |  | 0.7 | 11 | 94 | 0.97 |
|  |  | 0.8 | 12 | 98 | 0.98 |
|  |  | >0.8 | 14 |  |  |

BNP indicates B-type natriuretic peptide; DBP, diastolic blood pressure; LDL, low-density
lipoprotein; LVESD, left ventricular end-systolic diameter; LVMI, left ventricular mass index; RWT,
relative wall thickness; and SBP, systolic blood pressure.

## Discussion

We found that, for individuals without obstructive CAD, a simple comprehensive 10-variable scoring model (sex, smoke, SBP, DBP, BNP, LDL, LVMI, RWTLVESD, e’(l) and Tei) based on a combination of clinical, echocardiographic data and blood tests could offer a non-invasive means of identifying patients at earlier stages of developing CAS. The probability of CAS was <25% when the total score was ≤45; hence, it was reasonable to follow up the patients without performing coronary catheterization. A total score of ≥58 with the probability of CAS exceeding 50% indicates a significant chance of CAS being present and the specific non-invasive provocative hyperventilation testing should be considered. For those with a total score ≥69 with a very high probability of CAS ≥75%, coronary catheterization with CAS provocation tests is strongly recommended for a definite diagnosis. These results support the hypothesis that LV concentric remodeling and LV diastolic dysfunction may contribute to CAS. Our well-calibrated CAS risk score model had good discrimination, which could be helpful in prioritizing risks to allocate diagnostic coronary catheterization effectively and guiding treatment strategies.

While risk assessment is a critical step in primary prevention of cardiovascular disease, risk scores can be applied to stratify a population for targeted diagnosis. It has been well known that men are more likely than women to have epicardial CAS both in East Asia and Western countries.^5,36^ Men are known to have a higher prevalence and greater extent of coronary plaque burden than women, particularly in younger age groups,^37^ which, by using intracoronary imaging studies, may be associated with epicardial CAS.^38^ Furthermore, in CAS with minimal atherosclerosis, the survival rates are as high as 99% at 1 year and 94% at 5 years, while the survival in CAS with nonobstructive multivessel atherosclerosis falls to 87% and 77% at 1 and 5 years, respectively.^39^ Besides, even when compared with CAD patients, the incidence of cigarette smoking in men is significantly higher in the CAS than in the CAD group.^40^ In addition, sex differences in coronary physiology have been demonstrated by invasive and noninvasive studies of coronary flow. Middle-aged women compared to men exhibit more elevated resting coronary flow,^41^ likely related to sex differences in cardiac autonomic regulation.^42^ Previous invasive studies revealed that higher resting coronary flow may explain lower coronary flow reserve in women compared with men,^43,44^ albeit noninvasive studies suggest similar coronary flow reserve distribution in women and men.^45^ Moreover, premenopausal women have nearly 2-fold better coronary flow response than postmenopausal women and age-matched men,^46^ suggesting an important role of estradiol-mediated vasodilation.^47^ For a given epicardial stenosis severity, higher fractional flow reserve, another coronary physiological measure of an epicardial coronary stenosis, in women than in men could be due to sex-related differences in vasomotion,^48^ although women and men have similar index of microvascular resistance in ischemia with nonobstructive coronary artery disease.^43^ Taken together, the male predominance in atherosclerosis along with high smoking rate, lower resting coronary flow in men, the lack of estradiol-mediated vasodilation and lower fractional flow reserve might explain the higher prevalence of men than women in CAS development.

Smoking can lead to CAS through several mechanisms. First, the positive feedback loop between CRP-induced rise in patients’ monocytic IL-6 expression and nicotine-mediated α7-nicotinic acetylcholine receptors activation positively modulates CAS development.^49^ Second, smoking induces diffuse or segmental CAS, probably through adrenergic stimulation, catecholamine release, such as epinephrine, endothelial dysfunction^50^ and oxidative stress.^49^ Third, nicotine worsens coronary vasomotor dysfunction by platelet activation and raising blood viscosity, perhaps favored by endothelial dysfunction, may be a trigger of CAS.^51^ Collectively, smoking induces both direct nicotine’s chemical effects and indirect effects, including adrenergic stimulation, oxidative stress, inflammation, platelet activation and endothelial damage, that lead to CAS development.

While hypertension is found more frequently in classic angina than in CAS-induced angina,^52^ consistent with our results, Sugiishi et al. and Chen te al. found that low blood pressure was positively associated with CAS.^53,54^ Yasue et al. also revealed that the predictors for CAS were smoking, CRP and low DBP.^55^ We previously demonstrated that non-hypertensive smoking men and women than hypertensive smoking men (OR2.38, 95% CI 0.995–5.691) and women were at a higher risk for CAS in that order, respectively, with hypertensive non-smoking women being at the lowest risk.^5^ In rat aorta, hypertension is associated with premature ageing of the contraction of vascular smooth muscle cells in rat aorta.^56^ The contractile responses of coronary arteries to an inducer of CAS, serotonin, increase with age but are decreased by hypertension in rat coronary arteries.^57^ While synthetic phenotype smooth muscle cells are the main cultured vascular smooth muscle cell type from spontaneously hypertensive rats,^58^ contractile rather than synthetic phenotype smooth muscle cells play a main role in the pathogenesis of CAS. In addition, a drop in overall blood pressure, especially DBP, can directly cause temporary myocardial oxygen supply-demand imbalance and reduce coronary perfusion pressure, which would tend to potentiate the constrictor effects of endothelin; hence, endothelin during low blood pressure could be responsible for CAS.^59^ These apparently paradoxical findings collectively suggest that the pathogenesis of CAS differs from that of coronary atherosclerosis.

In 1956, two distinct experiments verified the heart as an endocrine organ, which decades later established the natriuretic peptide system that produces natriuresis and diuresis thirtyfold and tenfold, respectively, inhibits the renin-angiotensin-aldosterone axis, and acts as vasodilators.^60^ Likewise, that myocardial ischemia itself might activate similar effector loops is a novel concept. While elevated BNP levels can indicate the severity and impact of obstructive CAD,^61^ our results are in line with a previous study that BNP levels were lower in CAS-induced than in CAD-induced acute coronary syndrome,^11^ demonstrating the usefulness of BNP in differentiating a non-CAS-related from CAS-induced angina in non-obstructive CAD patients. On the other hand, intravenous infusions of synthetic BNP, such as nesiritide,^60^ are effective in suppressing hyperventilation-induced CAS^62^ as well as inducing coronary vasodilation in conductance and resistance arteries and suppress CAS.^60^ Furthermore, after pretreatment with endothelin to induce CAS, BNP increased coronary blood flow substantially and reversed endothelin-mediated vasoconstriction completely.^10^ These findings suggest that the coronary vasodilator effects of synthetic BNP and the longer half-life than atrial natriuretic peptide might favor BNP as a future option to prevent or treat CAS. Further studies in humans are required to examine the applicability of our findings to pathophysiological states in CAS. In line with a previous study showing LDL appeared to be a risk factor for CAS,^63^ we further demonstrated that the increased LDL is a risk factor of CAS. LDL is linked to CAS, particularly when small, dense LDL particles are involved.^64^ These particles are more prone to oxidation, and oxidized LDLs impair endothelial function and reduce the production of nitric oxide in CAS.^63^ Altogether, LDL, especially when it is oxidized, is linked to inflammation, endothelial dysfunction, and can contribute to CAS.

Despite ischemia with no obstructive CAD (INOCA), including CAS and coronary microvascular dysfunction, is increasingly recognized, they remine underdiagnosed and undertreated due to overreliance on global ejection fraction or fractional shortening, which is often normal even in patients with ischemic symptoms. Therefore, more sensitive imaging modalities for diastolic function are needed. The vasodilatory capacity of epicardial coronary vessels, which directly influences luminal volume, is regulated by both endothelial-dependent and -independent mechanisms.^65^ In a cohort of INOCA patients, coronary lumen volume to myocardial mass ratio was significantly lower compared to matched controls (25.6 ± 5.9 vs. 30.0 ± 6.5, p<0.001),^66^ emphasizing that the vasodilatory capacity of the epicardial vessels and the associated coronary lumen volume is potentially related to LV myocardial mass. As concentric remodeling determines LV diastolic dysfunction, which occurs early in the ischemic cascade, concentric remodeling confers increased cardiovascular risk compared to normal geometry^67^ and even those with eccentric LV hypertrophy.^68^ In the LIFE study, concentric remodeling was associated with a three- and eight-times increased risk of stroke and cardiovascular death after 4.8 years of follow up, respectively.^69^ From WISE study, in women with intermediate coronary flow reserve, those with lower myocardial perfusion reserve tended to have concentric remodeling and impaired LV diastolic function compared with those with higher myocardial perfusion reserve, providing evidence that both coronary vasomotion and concentric remodeling may be contributors to myocardial ischemia.^70,71^ On the other hand, in INOCA, impaired vasomotion, LV concentric remodeling, progressive fibrosis from repetitive coronary ischemia and the subsequent reduced myocardial capillary density may cause the consequent coronary flow reserve reduction, initiating increased metabolic demand caused by pressure overload and increased wall thickness.^71^ Our multivariable analysis revealed that, as a risk factor, RWTLVESD was more important than RWTLVEDD, suggesting that, along with impaired relaxation, CAS could reduce LV compliance and increase LV filling pressure. Regarding the relation to concentric remodeling in our CAS patients, mitral annular e’ velocity was negatively related to RWTLVEDD (r = −0.199, p<0.001), RWTLVESD (r = −0.152, p<0.001) and Tei index (r = −0.316, p<0.001) but positively related to E wave velocity (r = 0.330, p<0.001). Although our CAS and control patients had significantly different LVMI, they were within normal range; hence, the rarity of hypertrophy strongly suggests impairments in relaxation, rather than compliance per se, in CAS-induced ischemia was an important direct cause of diastolic dysfunction. Notably, in the WISE study, LV diastolic echocardiographic parameters, such as mitral annular velocities and Tei index, were not investigated and limiting their applicability.^70,71^ In these contexts, additional information to assist clinical decision-making is necessary. In women with INOCA, both the rate of diastolic strain and ventricular untwisting, reflected by the early doppler myocardial tissue velocities and the IVRT,^70^ respectively, are reduced than in controls, such that these abnormalities would presumably result in reduced e’(l) and e’(m) as well as prolonged IVRT, which are in line with our findings. This is also in accordance with previous findings demonstrating that a reduced diastolic function as determined by a reduced early diastolic relaxation tissue velocity e′ is a strong predictor of acute coronary syndrome.^72^ We demonstrated for the first time that CAS was associated with concentric remodeling, which may cause myocardial capillary rarefaction worsening diastolic function. Hence, probable positive interaction and modulatory loop may exist between CAS, diastolic dysfunction and concentric remodeling.

The Tei index is a parameter combining systolic and diastolic time intervals to assess global myocardial performance, which is reliable, reproducible, independent of heart rate and/or loading conditions. In cardio-oncology conditions, while 5-fluorouracil administration can induce CAS, post-infusion echocardiography reveals worsening of the Tei index as a sensitive indicator of silent 5-fluorouracil cardiotoxicity.^73^ In addition, since Tei index performs independently of preload and afterload, it may prove advantageous over the conventional load-dependent echocardiographic parameters that are routinely assessed. Notably, previous studies have demonstrated its good correlation with invasive measurements of cardiac function, including an inverse relationship with cardiac output, as well as direct correlation with systolic peak dP/dT, diastolic peak dP/dT, and ventricular stiffness.^74^ Since it is shown to be relatively load-independent, it could be especially useful to evaluate remodeled hearts, in which the altered preload and afterload conditions are likely to interfere with the standard echo parameters used to assess cardiac function, including ejection and shortening fractions. We further demonstrated that in CAS, Tei index was negatively related to mitral E wave velocity (r = -0.388, p<0.001) and BNP (r = -0.128, p=0.001) but positively related to DBP (r = 0.103, p=0.009), τ_0l_ (r = 0.208, p<0.001) and τ_0m_ (r = 0.202, p<0.001), while no correlation was found between Tei index and heart rate (r = 0.075, p=0.057). Tei index has been demonstrated significantly correlated with the diastolic peak (-dP/dt) and tau in people with ischemic heart disease;^74^ hence, its applicability is reasonably reliable for evaluation of LV function in CAS.

However, some limitations need to be acknowledged. First, the subjects in the study were patients in Shuang Ho Hospital, a reference center due to the quality of its interventional care, especially coronary angiography with CAS provocation test and myocardial revascularization. This could be a source of selection bias, but the data on validation of the risk scores reveals that the study sample included the full spectrum of CAS. Second, all risk estimation systems will underestimate risk if cardiovascular events have increased. Therefore, recalibration should be undertaken if good quality, contemporary event and risk factor prevalence data are available. Third, the findings of this study were derived from a single-center dataset consisting of an Asian population; therefore, the generalizability to other populations may be limited. Further studies involving diverse populations from different countries or races are warranted for recalibration and external validation.

## Conclusion

This hypothesis-generating study provides a simple comprehensive 10-variable scoring model (sex, smoke, SBP, DBP, BNP, LDL, LVMI, RWTLVESD, e’(l) and Tei) based on a combination of clinical, echocardiographic data and blood tests as a non-invasive means of identifying patients at early stages of developing CAS. When a total score of ≥58 with the predicted risk of CAS exceeding 50% was present, a high likelihood of CAS existed, and the specific non-invasive provocative hyperventilation testing should be contemplated. For those with a total score ≥69 with a very high probability of CAS ≥75%, coronary catheterization with CAS provocation tests is strongly recommended for a definite diagnosis. Our results indicated that LV concentric remodeling and LV diastolic dysfunction may contribute to CAS, encouraging insight into the pathophysiology of CAS and future studies to explore specific mechanisms as well as potential treatment options. Our well-calibrated CAS risk score model had good discrimination, which allowed for early allocating diagnostic coronary catheterization effectively, individualized treatments, and better outcomes.

## Data Availability

The data cannot be shared openly to protect study participants or patient privacy. However, the data supporting this study are available from the authors upon reasonable request.

## Non-standard Abbreviations and Acronyms

AUC: area under the curve
BNP: B-type natriuretic peptide
CAD: coronary artery disease
CAS: coronary artery spasm
CRP: C-reactive protein
hs-CRP: high sensitivity C-reactive protein
INOCA: ischemia with non-obstructive coronary artery disease
IVRT: isovolumic relaxation time
LDL: low-density lipoprotein
LVMI: left ventricular mass index
RWT: relative wall thickness
RWTLVEDD: relative wall thickness at end-diastole
RWTLVESD: relative wall thickness at end-systole.

## Acknowledgments

We thank Alfred Hsing-Fen Lin and Zoe Ya-Jhu Syu for their assistance in statistical analysis.

We also thank the intellectual support from Taiwan Society of Coronary Artery Spasm.

## Sources of Funding

This work was supported by National Science Council of Taiwan (NSTC 112-2314-B-038-104-MY3) and Taipei Medical University (114TMU-SHH-13) to Ming-Yow Hung.

## Disclosures

None.

## Notes

### Competing Interest Statement

The authors have declared no competing interest.

### Clinical Trial

Our study is not a clinical trial.

### Author Declarations

This study was approved by the Taipei Medical University-Joint Institutional Review Board (approval number: 201011004) and all patients gave written informed consent.

